# HIV-associated sensory neuropathy continues to be a problem in individuals starting tenofovir-based antiretroviral treatment

**DOI:** 10.1101/19002220

**Authors:** Prinisha Pillay, Antonia L Wadley, Catherine L Cherry, Alan S Karstaedt, Peter R Kamerman

## Abstract

HIV-associated sensory neuropathy (HIV-SN) is a common and often painful neurological condition associated with HIV-infection and its treatment. However, data on the incidence of HIV-SN in neuropathy-free individuals initiating combination antiretroviral therapies (cART) that do not contain the neurotoxic agent stavudine are lacking. We investigated the six-month incidence of HIV-SN in ART naïve individuals initiating tenofovir (TDF)-based cART, and the clinical factors associated with the development of HIV-SN. 120 neuropathy-free and ART naïve individuals initiating cART at a single centre in Johannesburg, South Africa were enrolled. Participants were screened for HIV-SN at study enrolment and then approximately every two-months for a period of approximately six-months. Symptomatic HIV-SN was defined by the presence of at least one symptom (pain/burning, numbness, paraesthesias) and at least two clinical signs (reduced vibration sense, absent ankle reflexes or pin-prick hypoaesthesia). Asymptomatic HIV-SN required at least two clinical signs only. A total of 88% of the cohort completed three visits within the six-month period. Eleven individuals developed asymptomatic HIV-SN and nine developed symptomatic HIV-SN, giving a six-month cumulative incidence of neuropathy of 140 cases per 1000 patients (95% CI: 80 - 210) at an incidence rate of 0.37 (95% CI: 0.2 - 0.5) per person year. Increasing height and active tuberculosis (TB) disease were independently associated with the risk of developing HIV-SN (p < 0.05). We found that within the first six months of starting cART, incident SN persists in the post-stavudine era, but may be asymptomatic.

## Introduction

The prevalence of HIV infection is high in sub-Saharan Africa and is expected to stay high for the foreseeable future ^1^. The high prevalence of HIV infections persists despite reduced infection rates because of the wide-spread availability of combination antiretroviral therapies (cART), which improve survival ^2^. But, this improved survival means that the total burden of chronic complications of HIV infection may increase, and therefore, these complications need to be monitored and managed to mitigate the impact they may have on the lives of people living decades with HIV ^3,4^.

One of the most common chronic neurological complications of HIV-infection and its treatment with neurotoxic combination antiretroviral therapy (cART) is HIV-associated sensory neuropathy (HIV-SN). The neuropathy is often symptomatic, with pain being the salient feature ^5,6^, and these symptoms have a detrimental impact on quality of life and function ^6,7^. Unfortunately, there are no widely-available pharmacological therapies with proven efficacy in the management of painful HIV-SN ^8–10^, which means that prevention of symptomatic HIV-SN is a key strategy. To limit side effects such as HIV-SN, the World Health Organisation (WHO) firstly lowered the dose, and then removed, the neurotoxic antiretroviral drug stavudine from all first-line cART regimens globally ^11,12^. With the worldwide elimination of stavudine in all first-line regimens and its replacement with tenofovir (TDF), an agent with no known neurotoxicity, together with earlier initiation of cART, it has been anticipated that the incidence of HIV-SN will decrease.

Several African studies have investigated the incidence of HIV-SN and factors associated with its development ^13–20^. However, none of these studies assessed the incidence of HIV-SN in a cohort universally exposed only to TDF-based cART. Instead, these studies included individuals who had been exposed to several ART regimens (including stavudine) or who were on cART regimens that did not include TDF (globally, the most common cART initiation regimen). Furthermore, the assessments used within these studies to assess HIV-SN status are methodologically questionable. For example, one study only assessed symptoms ^15^, while other studies did not state the method used to screen for HIV-SN ^13,14,17^.

Consequently, we investigated the six-month incidence of HIV-SN, using methods recommended for the assessment and grading of peripheral neuropathy for research and clinical purposes ^21–23^, in ART naive South African people living with HIV/AIDS (PLWHA) and who were initiating TDF-based cART. We investigated the six-month period after starting cART because on older stavudine-based cART, the neuropathy typically developed soon after starting therapy. As a secondary objective, we also described the clinical, disease-related and demographic risk factors associated with HIV-SN development in this cohort.

## Methods and Materials

We approached all treatment-naïve adult PLWHA who were initiating TDF-based cART at the Lenasia South Community Health Hospital, Johannesburg, South Africa, to partake in the study. Recruitment was conducted from January 2016 to December 2016. Participants were followed up for a period of approximately six-months. Ethical approval for the study was obtained from the Human Research Ethics Committee (Medical) of the University of the Witwatersrand, South Africa, clearance number: M121018. Written, informed consent was obtained from all participants. An interpreter fluent in English and six indigenous languages explained the study rationale and procedures in the participant’s language of choice.

### Study inclusion criteria

All study participants had to be ≥ 18 years of age, have a confirmed HIV diagnosis, have never received any antiretroviral treatment (including monotherapy) for their HIV infection, be initiating TDF-based cART, and be free of signs of distal symmetrical polyneuropathy (see neurological assessment below), although isolated lower limb symptoms were permitted.

### Study exclusion criteria

We excluded all participants who did not meet the inclusion criteria, and/or who had the presence of medical conditions that affected sensation in, or assessment of, the feet and lower limbs (e.g. Kaposi’s sarcoma).

Eligible participants had the following demographic and anthropometric data recorded: sex, age, height, weight, ethnicity, date tested positive for HIV infection, other potential risk factors for neuropathy (diabetes, alcohol consumption, vitamin B_12_ deficiency, diabetes mellitus and treatment for active TB disease). All participants with confirmed active TB disease were treated with rifafour [a four-drug combination TB disease treatment that includes the following: ethambutol, pyrazinamide (PZA), Isoniazid (INH) and rifampicin]. In addition, the above-mentioned participants were also given pyridoxine as prophylactic treatment to minimize the development of HIV-SN. Participants who did not have a confirmed diagnosis of active TB disease, received prophylactic Isoniazid Preventative Therapy (IPT), which consisted of INH (5mg/kg/day) and pyridoxine (started at 25mg titrated up to 100mg daily) for six to nine months. Participants without active TB had the choice of starting IPT.

The following laboratory data (where available) were obtained from participants medical records: CD4 T-cell count at the time of HIV testing (baseline CD4 T-cell count), current CD4 T-cell count (could be the same at baseline CD4 T-cell count), HIV viral load, current Hepatitis B infection, and current syphilis infection. A venous blood sample was taken from each participant at each visit to assess HBA1c percentage (HBA1c > 7% was used to define the presence of diabetes mellitus) and vitamin B12 concentrations (vitamin B12 < 142pmol/L was used to define clinically relevant deficiency).

### Procedures

#### Neurological assessment

Participants were assessed for the presence of peripheral sensory neuropathy (SN) using an amended version of the Brief Peripheral Neuropathy Screen (BPNS), which included the original BPNS plus the assessment of sensitivity to pin-prick. In its original format, the BPNS is a validated screening tool for HIV-SN ^24^, and has been previously used in the South African population ^25,26^. The BPNS case definition is for symptomatic HIV-SN and requires the bilateral presence of at least one symptom (pain/burning, paraesthesias, numbness), and the bilateral presence of one clinical sign (vibration sense in great toe and ankle reflex testing). We added the assessment of pin-prick sensitivity, to the BPNS tool for the following reasons: i) the two tests in the BPNS assess large fibre pathology, while pin-prick sensitivity assesses small-fibre pathology, ii) pin-prick sensitivity has high specificity in the detection of HIV-SN ^27^, and iii) we wished to the increase the specificity of the assessment (at the cost of sensitivity) by increasing the number of bilateral clinical signs required by our case definition to two signs (see case definitions below).

A diagnosis of *symptomatic* HIV-associated sensory neuropathy (HIV-SN) was made based on the bilateral presence of at least two signs (decreased vibration sense in the great toe, absent ankle reflex or decreased pin-prick sensitivity) and one symptom (pain, paraesthesias or numbness) in the feet. A diagnosis of *asymptomatic* HIV-SN was made based on the bilateral presence of at least two signs, but no symptoms, in the feet. By using participants medical history, symptomatology, clinical assessment of nerve function, and the anatomical distribution of signs and symptoms, we met the recommendations of England and colleagues ^21^, Haanpaa and colleagues ^22^, and achieved a diagnostic certainty (for symptomatic SN) of ‘probable neuropathic pain’ based on the grading criteria of Finnerup and colleagues ^23^.

Vibration sense was assessed using a 128Hz tuning fork on the distal interphalangeal joint of the participant’s great toe. Vibration sense was recorded in both feet, with a recording of 10 seconds or less deemed abnormal ^24^. Ankle reflexes were assessed using a rubber reflex hammer, where the investigator firmly struck the achilles tendon to elicit a response. A positive ankle reflex response was indicated by the plantar flexion of the foot and a negative response was recorded when there was no flexion of the foot. The reflex test was performed three times to confirm the response. Pin-prick sensitivity was assessed using a size 4 ‘single use’ safety pin. The ‘sharp’ and ‘blunt’ ends of the safety pin were alternately placed on the dorsal surface of the participants great toes, and participants were asked to identify the sensation experienced as either ‘sharp’ or ‘blunt’. Identification of the ‘sharp’ stimulus as being ‘blunt’ or that it was not felt at all was deemed abnormal. If participants gave an incorrect answer, we then tested lateral to the point and then progressed proximally in order to identify the extent/borders of nerve dysfunction.

When symptoms were present, the anatomical distribution of the symptom(s) was(were) recorded and participants were asked to rate the severity of the symptom(s) on an 11-point numerical pain rating scale (NRS), anchored at 0 (no symptom experienced) to 10 (worst pain/or sensation imaginable).

### Blood samples

One blood sample (4ml EDTA tube) was drawn from each at baseline and at every scheduled visit. Samples were analysed for CD4 T-cell count, HIV viral load, HbA1c levels and Vitamin B12 levels.

### Timing of follow-up

Study participants were recruited over about a six-month period, and each participant was followed up for six months, with four scheduled visits with the investigator. Following recruitment and consent procedures, visits were scheduled at 0 months (Baseline; day of treatment initiation), 2 weeks, 2-months, 4-months and 6-months after starting cART (supplementary material 2, Figure S2). Participants were contacted telephonically a few days prior to their scheduled return to the hospital to confirm their appointment day and to remind them to meet with the investigator.

### Statistical analysis

All analysis scripts and the outputs from the scripts can be accessed on Figshare (reserved doi: 10.6084/m9.figshare.7856660), or can be cloned from GitHub (https://github.com/kamermanpr/hivsn-incidence.git).

Descriptive statistics are presented as mean (standard deviation, SD) for parametric data, median (interquartile range, IQR) for non-parametric data, and percentages for frequency data. 95% confidence intervals of the mean/median difference and odds ratios between ‘SN-free’ individuals and individuals with SN were computed for analysis of factors associated with SN across the study period.

Our primary objective was to determine the incidence of HIV-SN. We used both cumulative incidence data and incidence rate data to determine the incidence of HIV-SN in our cohort. We first determined cumulative incidence, which measures the number of new cases per person in the population over a defined period of time (i.e. a fixed follow-up period). Therefore, to calculate cumulative incidence we defined a fixed 6-month follow-up period. We also subdivided this period into a 1st and 2nd 3-month period of this follow-up. To standardize the periods of follow-up, we cleaned the data and used only visits that fell into the indicated periods: (0-3 months), (3-6 months) and (0-6 months) to define SN status. Using this method, participant’s whose last clinic visit occurred after 91 days (end of first 3-month interval) or at 182 days (end of 6-month interval), and who were found to have new-onset SN at this visit, were recorded as ‘SN-free’ over the 3 or 6-month period of follow-up, respectively. This conservative strategy may have led to some under-estimation of the cumulative incidence of SN. We also determined incidence rate, which is a measure of the number of new cases per unit of time. We did not have exact dates of SN onset to define the per patient unit of time, and nor was there uniform spacing between clinic visits. We therefore chose to calculate an approximate SN onset time, arbitrarily defined as the number of days between the first neuropathy screening and the mid-point between the visit when neuropathy was detected and the preceding visit. For participants who did not develop SN, the date of censoring was defined by the number of days between the first neuropathy screening and the last screening (study exit).

Our secondary objective was to determine possible risk factors for developing HIV-SN in our cohort. We first carried out an exploratory analysis using Cox proportional hazard models, however, various predictors violated assumptions of the model (e.g., proportional hazard, linearity, no influence points). We therefore performed multivariable logistic regression analysis with elastic net regularization for variable selection, with visit 1 (baseline) characteristics as predictors of SN onset. Elastic net regularization performs both variable selection and regularization in order to enhance the prediction accuracy and interpretability of the statistical model it produces. The process involved performing a 10-fold cross-validation to find the optimal lambda (penalization parameter) and thereafter running the analysis and extracting the model based on the best *lambda*. The advantages of regularized regression methods come at the cost of biased estimates, and so we have not reported the odds ratios. The models included the baseline values of all variables, except diabetes mellitus (as no participants had diabetes at baseline), Vitamin B_12_ deficiency (as only one person had a deficiency at baseline) and weight as it was co-linear with height.

## Results

From January 2016 through to December 2016, 151 confirmed HIV-positive individuals were assessed for eligibility for the study. One hundred and twenty individuals met the requirements for inclusion in the study (Figure 1). Of the individuals not fulfilling the entry criteria, and so not recruited, nine (6%, 9/151) had a pre-existing neuropathy. All study participants identified themselves as of black African ancestry; 104 (87%) were South African.

**Figure 1.**
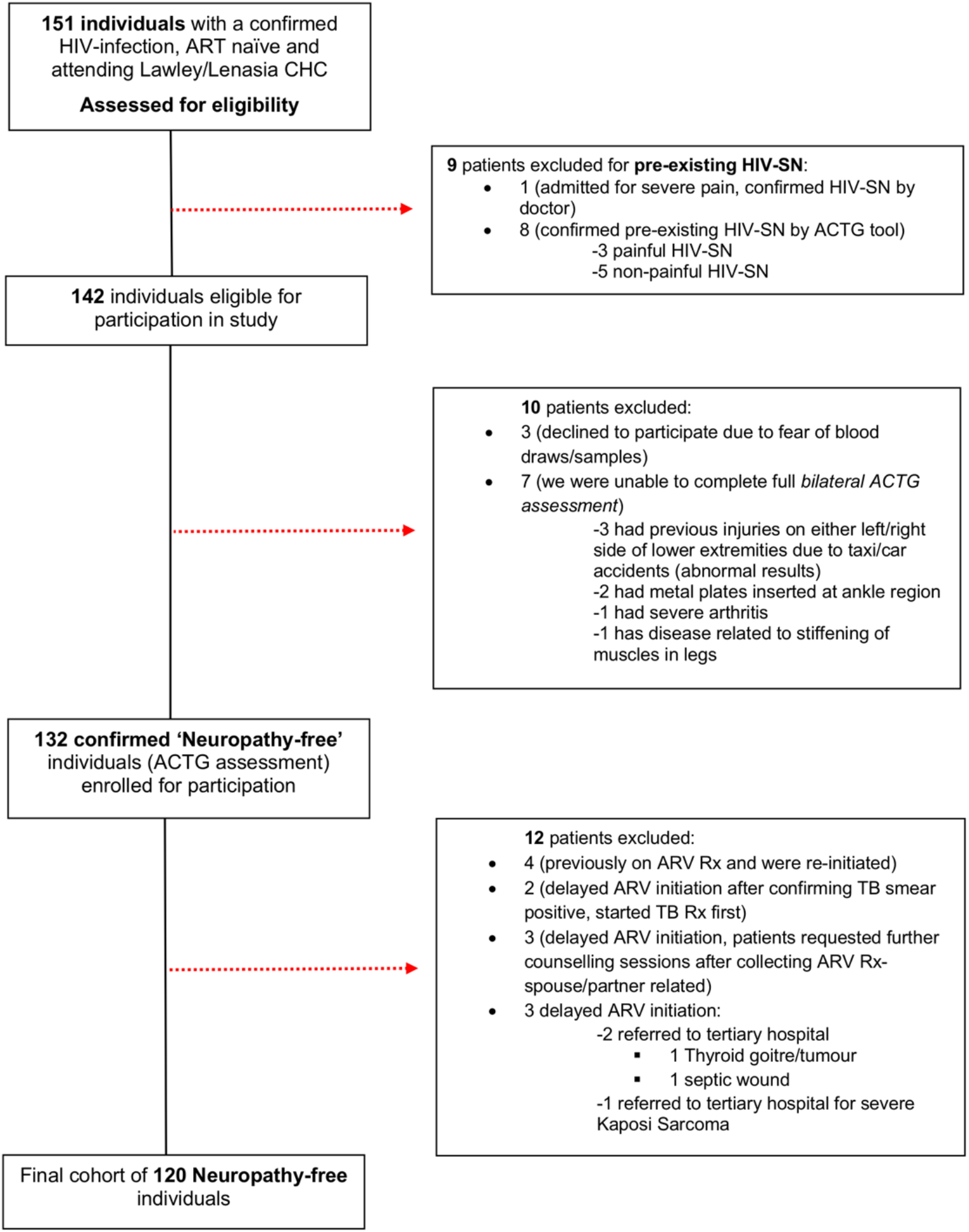
Flow diagram summarising participant recruitment and exclusion.

### Demographic and clinical characteristics

Baseline demographic and clinical characteristics of the study cohort are shown in Table 1. Over three quarters of the cohort were categorized under WHO stage III and IV (92/120, 77%), indicating that the majority of our patients were immunocompromised. The median (range) CD4 T-cell count at initiation of the study was 228 (1-1347) cells/µL. No participants had ever previously been exposed to any other antiretroviral therapy, and all participants were initiated on TDF-based fixed-dose triple combination [tenofovir + emtricitabine + efavirenz (TDF + FTC + EFV)]. One sixth of participants (20/120, 17%) were currently being treated for active TB disease with rifafour with pyridoxine (vitamin B6) prophylaxis. About one third of participants (41/120, 34%) without a diagnosis of active TB disease were being treated prophylactically with IPT.

**Table 1.** Demographic and clinical characteristics of the study cohort at baseline (n=120).

| Characteristic | Entire cohort (n=120) |
| --- | --- |
| Female Sex, n (%) | 66 (55) |
| Age (years) | 39 (12) |
| Height (m) | 1.63 (0.08) |
| Weight (kg) | 65.0 (15) |
| Current CD4 T-cell count (cells/μL) | 228 (1-1347) |
| log<sub>10</sub> viral load (copies/mL) | 3.19 (1.69-6.5) |
| WHO staging, n (%) |  |
| I | 13 (11) |
| II | 15 (12.5) |
| III | 37 (31) |
| IV | 55 (46) |
| Starting cART regimen, n (%): |  |
| *TDF-based* | 120 (100) |
| *Non TDF-based* | 0 (0) |
| Active TB disease, n (%) | 20 (17) |
| Standard units of alcohol consumed (units /week) | 15.0 (3-90) |
| Initial Vitamin B<sub>12</sub> level (pmol/L) | 316.5 (140-693) |
| Vitamin B<sub>12</sub> deficiency, n (%) | 1 (1) |
| Current hepatitis B infection, n (%) | 4 (3) |
| Current syphilis infection, n (%) | 3 (3) |
| Initial HbA1c percentage | 5.4 (4.0-7.0) |
| HbA1c > 7%, n (%) | 1 (1) |
| Language group, n (%) |  |
| *isiZulu* | 45 (38) |
| *isiXhosa* | 20 (17) |
| *Sesotho* | 26 (22) |
| *Setswana* | 7 (6) |
| *Other* | 22 (18)<sup>a</sup> |

<sup>a</sup>Other, n (%): Tsonga, 2 (2); Venda, 2 (2); Pedi, 3 (3); Afrikaans, 3 (3), Portuguese 7 (6), Chewa 4 (3), Shona 1 (1).
Dichotomous variables are presented as number (%), normally distributed continuous variables are presented as mean (standard deviation) and non-normally distributed continuous variables are presented as median (range).

**Table 2.** Elastic net variable selection.

|  | **Alpha<sup>†</sup>** |  |  |  |
| --- | --- | --- | --- | --- |
|  | **0.125** | **0.216** | **0.343** | **1.0** |
| Age (years) | + | + |  |  |
| Female sex (yes) |  |  |  |  |
| Height (m) | + | + | + | + |
| CD4 (cells/μl) | | | | |
| Viral load (copies/ml) |  |  |  |  |
| Alcohol (units/week) | + |  |  |  |
| TB current (yes) | + | + | + | + |
| Rifafour<sup>a</sup>/INH<sup>b</sup> (yes) |  |  |  |  |

† Alpha is the mixing parameter between L2 (ridge) and L1 (lasso) regularization.
<sup>a</sup> Participants with active TB disease received rifafour plus pyridoxine prophylaxis.
<sup>b</sup> Participants without active TB disease received isoniazid and pyridoxine, purely as prophylactic treatment.

### Symptom prevalence at baseline

At baseline, 20% of participants (24/120) presented with pain, 14% (17/120) had paraesthesias, and 5% (6/120) presented with numbness in the lower limbs. Despite the presence of these symptoms they were all classified as ‘neuropathy-free’, as none of these participants presented with any signs of SN. Symptoms were located primarily in the foot region for 85% (40/47) of participants with symptoms, with the remaining 15% (7/47) of participants experiencing the extension of these symptoms up to the ankle and knee regions. In participants with pain, the median pain intensity was 5 (IQR 2-10) on an 11-point numerical pain rating scale.

### Patient follow-up

Of the cohort, 88% had at least 3 clinic visits. Although participant follow-up was planned for 0 (the day of cART initiation), 2, 4 and 6-months of cART, there was variability in the visit schedules of participants as depicted in (supplementary material 3, Figure S3). Timing of clinic visits stratified according to SN status is provided in Figure 2.

**Figure 2.**
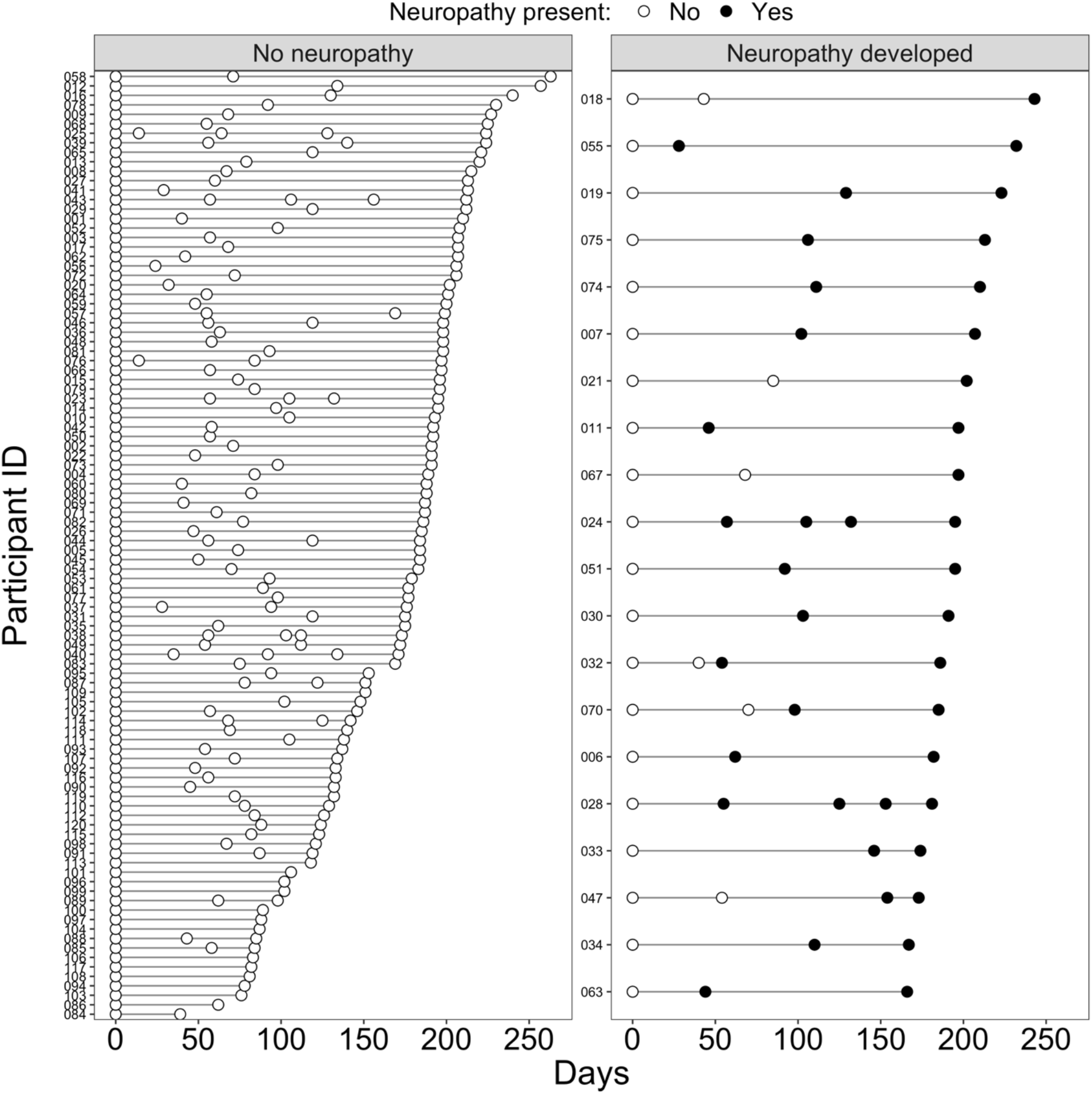
Timing of clinic visits for the entire cohort, stratified according to the development of HIV-SN. Open circles show visits where participants were ‘SN-free’, while closed circles show visits where participant were identified as having developed SN.

The median (IQR) number of clinic visits for the entire cohort (n=120) was 3 (3) (supplementary material 3, Figure S4). There was no difference in the number of clinic visits between those individuals that remained ‘SN-free’ [median (IQR): 3 (3-3)] and those that developed SN [3 (3 - 3.25)]; *X*^2^ (df:1) = 4.35, p = 0.2]. There was also no difference in the number of days between the first and last clinic visit between those individuals that remained ‘SN-free’ [median (IQR) 184 (133-199)] and those that developed SN [195 (182-208)], (Z = −2.10, p= 0.03; Two sample van der Waerden test) (supplementary material 3, Figure S5), or days between successive visits (F(1) = 0.02, p = 0.87; Type II Wald F test with Kenward-Roger df) [supplementary material 3, Figure S6].

### Incidence of neuropathy

Of the 120 confirmed ‘SN-free’ patients who were enrolled at baseline, a total of 31% (37/120) of patients were lost to follow-up by the 6-month study period. However, 88% (105/120) completed at least three visits or more. A total of 17% (20/120) developed HIV-SN and 83% (100/120) remained ‘SN-free’. Using these data, we calculated the cumulative six-month incidence of HIV-SN to be 140 cases per 1000 patients (95% CI: 80 - 210). The cumulative incidence over the first three-months of follow-up was 60 cases per 1000 patients (95% CI: 20 - 100), and for the second three-months of follow-up was 90 cases per 1000 patients (95% CI: 40 - 140). The incidence rate of SN was 0.37 (95% CI: 0.2 - 0.5) per person-year. Nine (8%) individuals developed symptomatic HIV-SN and eleven (9%) individuals developed asymptomatic HIV-SN.

The presence of symptoms at baseline (including pain, paraesthesias, numbness and burning) were associated with an increased risk of development of HIV-SN across the six-month period [*X*^2^ (df:1) = 15.09, p< 0.001]. Of the (47/120, 39%) of participants that had presented with symptoms at baseline, 13% (6/47) developed symptomatic HIV-SN and 9% (4/47) developed asymptomatic HIV-SN. The median pain intensity of the 15% (3/20) patients that developed symptomatic HIV-SN in the first three-month period, was 4 (IQR: 3-5). The median pain intensity of the 30% (6/20) patients that developed symptomatic HIV-SN in the second three-month period, was 5 (IQR: 4-10).

#### Survival curve analysis

Figure 3 shows the Kaplan-Meier survival curve with the number and percentage of patients at risk for developing neuropathy across the study period. Of the 20 individuals that developed neuropathy, 7% (8/120) of patients developed neuropathy within the first 50 days of cART; one patient developed neuropathy at about two weeks. Within the next 100 days of cART, 10% (12/120) individuals developed neuropathy. The last event of neuropathy development occurred at about 140 days. A secondary Kaplan-Meier survival analysis of whether the onset of neuropathy differed between those who developed a painful neuropathy compared to those with an asymptomatic neuropathy found that the onset of painful neuropathy was more rapid than the onset of asymptomatic neuropathy (*X*^2^ (df:1) = 13.4, p< 0.001; supplementary material 4, Figure S7). The first painful neuropathy occurred at ± two weeks. The development of asymptomatic peripheral sensory neuropathy had a slightly delayed onset with the first asymptomatic neuropathy occurring at about day 25.

**Figure 3.**
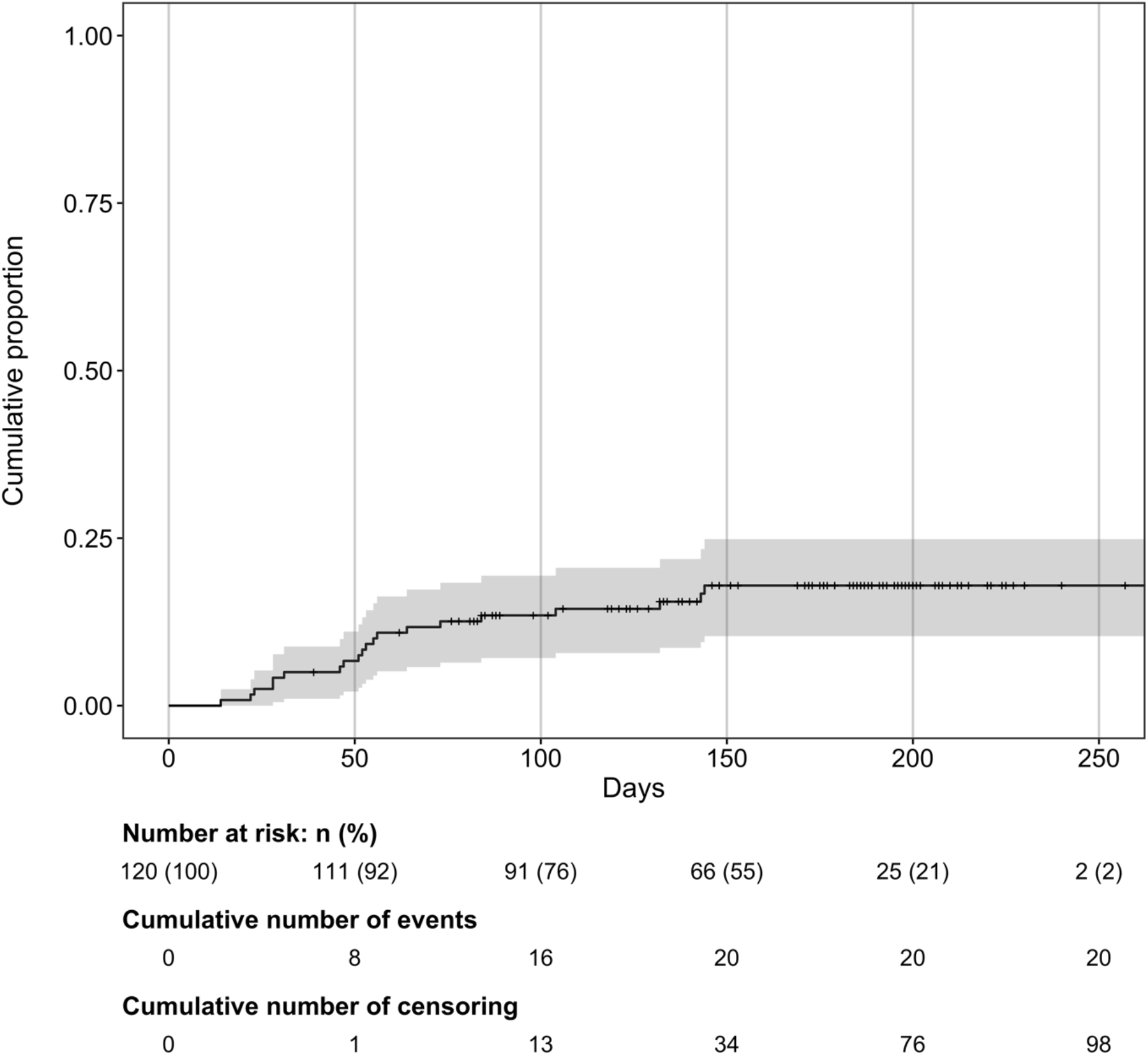
Kaplan-Meier survival curve showing the number and percentage of participants at risk for developing neuropathy across the study period.

### Multivariable analysis

We included baseline clinical, demographic, and physical factors in the modelling process. The full model included: sex, active TB disease, age, rifafour/INH prophylaxis treatment, CD4 T-cell count, height, viral load, and alcohol consumption. Across all alphas, and the optimal lambda at each alpha, the models repeatedly returned only two variables: increasing height and active TB disease.

## Discussion

We investigated the six-month incidence of HIV-SN in a cohort of PLWHA of black African ancestry who were universally exposed to TDF-based cART. We report a six-month cumulative incidence of HIV-SN of 140 cases per 1000 patients (95% CI: 80 - 210). The incidence rate was 0.37 (95% CI: 0.2 - 0.5) per person year. A roughly equal number of individuals developed symptomatic (8%, 9/120) vs. asymptomatic (9%, 11/120) SN. The presence of neuropathic pain was associated with a more rapid onset of symptomatic HIV-SN. Increasing height and having current active TB disease were the only consistent independent predictors of HIV-SN in the cohort across several modelling parameters. Given the absence of any neurotoxicity by TDF-based regimens, we do not believe that the continued occurrence of HIV-SN is a direct effect of the treatment regimen on peripheral nerves.

Our findings are novel and provide the first incidence data for individuals solely exposed to TDF-based cART. Our findings indicate that the number of new cases of HIV-SN is still high, even if substantially lower than those observed in a similar cohort initiating stavudine-based cART ^28^. In the previously mentioned cohort, 44 individuals initiated stavudine-based therapy, and 41% (18/44) of these individuals developed SN within 6-months ^28^. Our findings in the era of TDF-based cART highlight two key facts: i) Incidence of HIV-SN is lower on TDF-based cART, but still higher than we anticipated based on the lack of neurotoxicity of these drugs, and ii) HIV-SN in the TDF-based era tends to present asymptomatically more often compared to stavudine-based therapy where symptomatic HIV-SN was more prevalent [∼61%] ^28^.

While the increase in the proportion with asymptomatic neuropathy is welcomed, this finding needs to be regarded with some caution: we only followed individuals for six months and so it is not clear if asymptomatic SN remained asymptomatic or whether it may have progressed to being symptomatic. Indeed, one high-quality study suggested that asymptomatic SN may transition into symptomatic HIV-SN within a 12 to 24-month period on cART ^29^. When symptomatic neuropathies developed within our cohort, they developed rapidly (most cases occurring within 3-months of initiating cART, and as early as two weeks), compared to asymptomatic neuropathies, which developed more slowly (most cases within ± 5-months of initiating cART). Rapid onset, painful neuropathies have also been reported in cohorts on other cART regimens including stavudine, zidovudine and zalcitabine ^30^.

The data regarding associations between HIV-SN and disease markers such as CD4 T-cell count and viral load are equivocal ^31–33^. Indeed, we found no association with these variables and HIV-SN. However, a role for immune dysfunction in mediating HIV-SN cannot be excluded. For example, animal models of HIV and ART-induced neuropathy show immune activation in the dorsal root ganglia and nerve trunks, and these changes mimic those seen in post-mortem studies in humans with HIV-SN (for review: ^34^. Moreover, there are preliminary data suggesting a particular inflammatory profile may be responsible for neuropathy, and particularly, rapid onset painful neuropathy. In individuals starting cART, patients with greater interleukin-1-receptor (IL-1R)-antagonist levels at baseline, and two weeks later elevated levels of soluble interleukin-2 receptor-alpha and tumour necrosis factor (TNF) receptor-II were more likely to develop a symptomatic HIV-SN ^19^. Thus, it is possible that a particular inflammatory response results in a faster onset and painful HIV-SN, rather than exposure to a particular ART-regimen. Future studies are required to compare inflammatory profiles between those developing fast and slow onset HIV-SN.

The presence of active TB disease was an independent predictor of HIV-SN in our cohort. Importantly, there was no association found between rifafour exposure and HIV-SN, in our setting. The association between HIV-SN and treatment for active TB disease is well documented on stavudine-based regimens, even with pyridoxine supplementation ^18,25,35–37^. However, our study is the first study to associate TDF-based cART and confirmed active TB disease with the development of HIV-SN. The association between active TB disease and HIV-SN in those individuals starting ART, may not be specific to the type of ART, therefore, but perhaps related to the timing of ART and concurrent active TB disease. The timing of ART initiation in the face of active TB disease can lead to TB-immune reconstitution inflammatory syndrome (IRIS). This condition involves a strong inflammatory response to TB antigens during the acute phase of immune system reconstitution that leads to clinical deterioration ^38^. If such immune dysregulation also is associated with immune activation in peripheral nerves, then the probability of clinically apparent neuropathy would increase. However, none of our participants developed frank TB-IRIS, so this cannot be the mechanism underlying this association. Instead, we hypothesize concurrent active TB disease may reduce the decrease in inflammation that typifies the initiation of cART ^39^, thus predisposing affected individuals to develop a neuropathy. Whatever the mechanism, if the relationship between HIV-SN, concurrent active TB disease and cART is causal, then HIV-SN will remain a problem in countries with high co-infection rates despite the introduction of non-neurotoxic regimens.

Increased height was an independent predictor of HIV-SN in our cohort. Increasing height is a well-established risk factor for length-dependent polyneuropathies such as HIV-SN and diabetic sensory polyneuropathy ^40^. Therefore, it is not surprising that taller individuals would be more susceptible to the development of HIV-SN ^41^.

There are several limitations to our work. One of these limitations was the sample size of our cohort. We aimed to recruit a sample size of n=150 individuals, although we did not achieve this number of participants, our cohort still consists of the largest study of HIV-SN in ART naïve individuals initiating TDF-based cART. Our sample size was a direct result of the referral of ART naïve individuals to larger tertiary-based hospitals when they presented to the local clinic (where our recruitment was based) with existing opportunistic infections such as cryptococcal meningitis, pneumonia and Kaposi’s’ sarcoma. These opportunistic infections are better managed in these large tertiary hospitals, due to the availability of specialist care. Such individuals were initiated on ART at these tertiary hospitals and they typically did not return to their local clinic, thereby reducing the number of ART naïve individuals available at site for study recruitment. It also meant that our cohort represented uncomplicated cases of HIV infection, and our results cannot be extrapolated to more complicated cases. The other limitation was our retention of participants. We aimed to retain as many study participants as possible across the six-month period of our study. Despite having specific times for patient visits on our protocol and allowing ± 2 weeks before or after scheduled visits for patients to come in for their follow up visits, we faced several issues at the research site which were out of our control and which disrupted the timing of participants returning for follow-up visits. These issues included frequent community service delivery protests at our research site, which were violent and resulted in all entrances to the hospital being blocked.

## Conclusion

We report that the incidence of HIV-SN on TDF-based cART is indeed lower than that observed on stavudine-based therapies, and HIV-SN that develops in TDF-exposed individuals is more likely to be asymptomatic. Whilst these findings are good news, it is important not to become complacent. There was still a percentage of individuals who developed a rapid onset symptomatic neuropathy, which we know will impact quality of life irrespective of the severity of the symptoms ^42^. Active TB disease associated with development of HIV-SN in the cohort here. HIV-SN may well continue to be a clinical problem in resource-poor countries in Africa and India where prevalence of comorbid TB and HIV remains high.

## Data Availability

Study participants did not consent to the public release of their data. However, the data are available on request from Peter Kamerman.

https://github.com/kamermanpr/hivsn-incidence.git

## Acknowledgments

The authors thank the patients and staff of the Lenasia South Community Health Hospital (CHC), and Florence Mtsweni for acting as an interpreter. We thank Miss Onkarabile Seane of the Chronic clinic within the Lenasia South Community Health Hospital for her assistance. Medical Faculty Research Endowment Fund of the University of the Witwatersrand (PP). Medical Research Council of South Africa (PRK). National Research Foundation Rated Researchers Programme (PRK). The authors gratefully acknowledge the contributions of the Victorian Operational Infrastructure Support Program received by the Burnet Institute (CLC)

